# Clinical characteristics of COVID-19 patients in Latvia under low incidence in Spring 2020

**DOI:** 10.1101/2020.12.22.20239392

**Authors:** Ludmila Viksna, Oksana Kolesova, Aleksandrs Kolesovs, Ieva Vanaga, Seda Arutjunana, Sniedze Laivacuma, Jelena Storozenko, Ieva Tolmane, Ilze Berzina, Baiba Rozentale

**Affiliations:** Departments of Infectology, Rīga Stradiņš University, Latvia; Joint Laboratory of Immunology and Immunogenetics, Rīga Stradiņš University, Latvia; Faculty of Education, Psychology, and Art, University of Latvia, Riga, Latvia; Faculty of Medicine, University of Latvia, Riga, Latvia; Department of Public Health and Epidemiology, Rīga Stradiņš University, Latvia; Riga East Clinical University Hospital, Riga, Latvia

**Keywords:** COVID-19, blood tests, severity, hospitalization, tissue damage, renal function

## Abstract

**Background:** COVID-19 is a new infectious disease with severe disease course and high mortality in some groups. Blood tests on admission to the hospital can be useful for stratification of patients and timely correction. Our study investigated the clinical features of COVID-19 patients in Latvia and differences in blood tests in groups with different disease severity.

**Methods:** The retrospective study included 100 patients hospitalized in Riga East Clinical University Hospital in Spring 2020. The severity of the disease course was classified by the presence of pneumonia and its combination with respiratory failure. We have assessed blood cells’ count, hemoglobin, hematocrit, erythrocyte sedimentation rate (ESR), C-reactive protein (CRP), alanine aminotransferase, lactate dehydrogenase (LDH), troponin T, electrolytes, creatinine, glomerular filtration rate (GFR), D-dimer, prothrombin time, prothrombin index, oxygen saturation, and temperature on admission to the hospital.

**Results:** Patients were from 18 to 99 (57±18 years, 57% males). Comorbidities were found in 74% of patients. The mild, moderate, and severe groups included 35, 44, and 16 patients, respectively. In the severe group, the mortality rate was 50%. The progression to severe COVID-19 was associated positively with temperature, ESR, CRP, creatinine, LDH, and troponin T and negatively associated with oxygen saturation, eosinophils, and GFR on admission to the hospital.

**Conclusions:** COVID-19 severity associates with lower renal function and a higher level of inflammation and tissue damage. Eosinophils, CRP, ESR, LDH, troponin T, creatinine, and GFR are blood indicators for monitoring patients’ condition.

## Introduction

Coronavirus Disease (COVID-19) is a new infectious disease with a variable course, multiple organ dysfunction in severe cases, and mortality risk [1]. In the middle of December, there were 75,704,061 confirmed cases and 1,690,061death globally [2]. A meta-analysis of retrospective studies [3] demonstrated that severe COVID-19 disease course is observed in 12.6% to 23.5% of patients, while in groups with comorbidities, it was up to 44.5%. The mortality range is from 2.0% to 4.4 %, but it can reach 41.7% in patients with diabetes and hypertension.

Routine blood tests can help clinicians to assess the severity and prognosis of patients with COVID-19 [4]. Previous studies on clinical characteristics of patients with COVID-19 described that disease severity associates with low lymphocyte count on admission, comorbidities, and gender [5, 6, 7]. Moreover, C-reactive protein (CRP), procalcitonin, D-dimer levels, and leukocyte count were significantly higher in severe than in non-severe cases of COVID-19 [8]. A meta-analysis of 6320 patients from 52 studies [9] has included 36 laboratory parameters showed that the progression to severe phenotype also associates with elevated levels of neutrophils, prolonged prothrombin time, fibrinogen, erythrocyte sedimentation rate, IL-6, and IL-10 on admission to the hospital. However, most of these studies were conducted in Asian countries. European countries were not presented in this analysis, and we have reacted to the need for locally relevant data on disease course severity.

Latvia is one of the Baltic region countries, which reported the lowest total COVID-19 cases and death than in neighboring Estonia, Lithuania, Belarus, and Russia throughout the pandemic period in Spring 2020. In Latvia, the first case of SARS-CoV-2 infection was reported on 3 March 2020. At that time, the SARS-CoV-2 infection was diagnosed in 670 people in Europe [10]. During the first three months of pandemics, 1,066 total cases and 25 deaths were registered in Latvia [10]. This relatively favorable situation in Spring 2020 was a reason for speculations on clinical features of COVID-19 patients in Latvia and their deviance from those of patients in other countries.

This study investigated the clinical features of COVID-19 patients hospitalized in the largest hospital of Latvia, providing treatment for most patients in the country. It aimed to reveal differences in blood tests in patients with different COVID-19 severity, which was classified by the presence of pneumonia and its combination with respiratory failure.

## Material and methods

The retrospective study started in July 2020. The Permission of the Central Medical Ethics Committee (protocol No. 01-29.1/2429) and Permission of the Ethics Committee of Riga Stradinš University (protocol No. 6-1/07/14) were obtained for this study. All patients were hospitalized in Riga East Clinical University Hospital (the largest hospital in Latvia) from March to June 2020. All patients had positive real-time reverse-transcription polymerase chain reaction (RT-PCT) for SARS-CoV-2 nucleic acids of a nasopharyngeal swab.

For laboratory tests, blood samples were received within 24 hours of admission to the hospital. Blood tests included the following groups of parameters:

*Blood cells:* white cells count (WBC), neutrophils (Neu), lymphocytes (Ly), eosinophils (Eo) and monocytes (Mo), erythrocytes (RBC), platelet count (PLT), hemoglobin (Hb), and hematocrit (HCT);

*Inflammatory indicators:* erythrocyte sedimentation rate (ESR) and C-reactive protein (CRP);

*Tissue damage indicators:* alanine aminotransferase (ALT), lactate dehydrogenase (LDH), and troponin T (TnT);

*Electrolytes:* potassium and sodium concentration;

*Renal function indicators:* creatinine and glomerular filtration rate (GFR);

*Coagulation tests:* D-dimer, prothrombin time, and prothrombin index.

For analysis, also data of body’s temperature and SpO_2_ on admission were used.

For detection of the clinical course of COVID-19 medical documents were used. The severity of the clinical course of COVID-19 was assessed by using data about radiologically confirmed pneumonia and the development of respiratory failure. Disease outcome was defined as discharge or death.

We have used IBM SPSS 22.0 for computations. For a comparison of three groups, the Kruskal-Wallis test was applied. Following multiple pairwise comparisons included the Dunn-Bonferroni significance correction. The Chi-square test was used for the assessment of the distribution of nominal variables (e.g., gender or outcome). The Spearman rank correlation coefficient was applied for the assessment of relationships among laboratory indicators.

## Results

In total, 100 patients were included in the study. Table 1 presents demographics, epidemiological history, comorbidities, and symptoms on admission to the hospital.

**Table 1.** Demographic, epidemiological, and clinical data of patients (n=100)

| Characteristics | Overall |
| --- | --- |
| **Demographics** |  |
| Age, (s.d.), years | 57 (18) |
| Males / females, % | 57 / 43 |
| Riga is place of residence, % | 66 |
| **Epidemiological history, %** |  |
| Return from travel two weeks before the illness | 26 |
| Contact with suspected patients | 63 |
| Contact with confirmed patients | 51 |
| **Comorbidities, %** | 74 |
| Arterial hypertension | 44 |
| Chronic heart failure | 14 |
| Atherosclerosis | 15 |
| Arrhythmias | 12 |
| Diabetes mellitus | 11 |
| Chronic obstructive pulmonary disease | 4 |
| Asthma | 8 |
| Chronic kidney disease | 6 |
| Adiposity | 8 |
| Cerebrovascular disease | 8 |
| Oncological disease | 5 |
| Liver diseases | 4 |
| **Medication before hospitalization, %** |  |
| Daily medication | 50 |
| Immunosuppressive therapy | 1 |
| **Symptoms on admission, %** |  |
| Fever | 77 |
| Temperature > 38.0 °C | 63 |
| Cough | 74 |
| Dry | 52 |
| Productive | 15 |
| Paroxysmal | 6 |
| Fatigue | 35 |
| Varying degrees of shortness of breath | 20 |
| Heaviness in the chest | 4 |
| Headaches | 10 |
| Muscle pain | 2 |
| Sore throat | 3 |
| Scratching in the throat | 2 |
| Gastrointestinal symptoms | 6 |
| Loss of smell or taste | 1 |
| Dizziness | 6 |
| Anxiety and fear | 4 |
| Skin rash | 1 |
| Pulse rate > 80 beats per minute | 62 |

The mean interval from the onset of symptoms to admission to the hospital for all patients was 7±5 days (0 to 24 days). The most common complaint on admission to the hospital was fever (77%). The mean body temperature on admission was 37.9±0.9 °C (min - 36.2 °C, max - 40.0 °C), 63% of patients had a temperature above 38.0 °C. The second symptom was cough (74%), more often dry cough. The third self-reported symptom was fatigue (35%). One-fourth of patients have mentioned varying degrees of shortness of breath. Other symptoms were relatively rare. Gastrointestinal symptoms such as diarrhea, nausea, vomiting, or loss of appetite were reported in 6% of patients.

The mean time of the hospitalization was 12±8 days (1 to 40 days). Fifty-nine percent of COVID-19 patients developed pneumonia during their illness, and 15% had respiratory failure. Acute liver damage during the COVID-19 was detected in 1% of patients and thromboembolism in 1%. Secondary bacterial infection was reported in 5% of patients.

In our study, all patients were categorized into three groups: mild, moderate, and severe cases. Mild cases included patients without X-ray confirmed pneumonia or respiratory failure. Moderate cases included patients with X-ray confirmed pneumonia and without respiratory failure. One patient from this group had admission to the Intensive Care Unit (ICU). Severe cases included patients with confirmed pneumonia and respiratory failure. Sixty-six percent of the severe group had admission to ICU.

The mild group included 35 patients with COVID-19 (median age 49). The group of moderate cases included 44 patients (median age – 65), and the group of severe cases included 16 patients (median age was 59). The mortality rate was 2.3% in the moderate group and 50% in the severe group that associated significantly with the disease course (Table 2). All patients of the mild group were discharged.

**Table 2.**
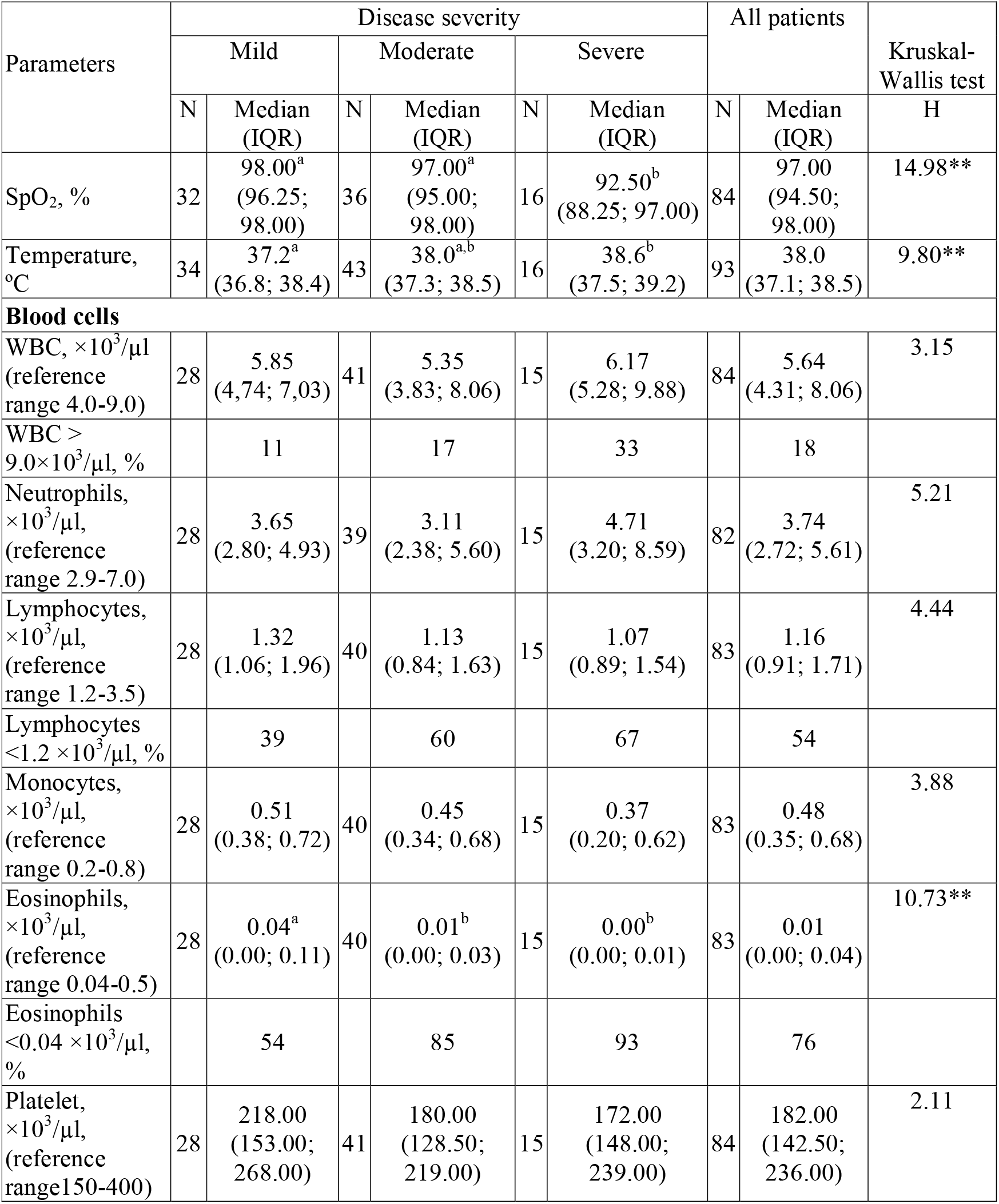
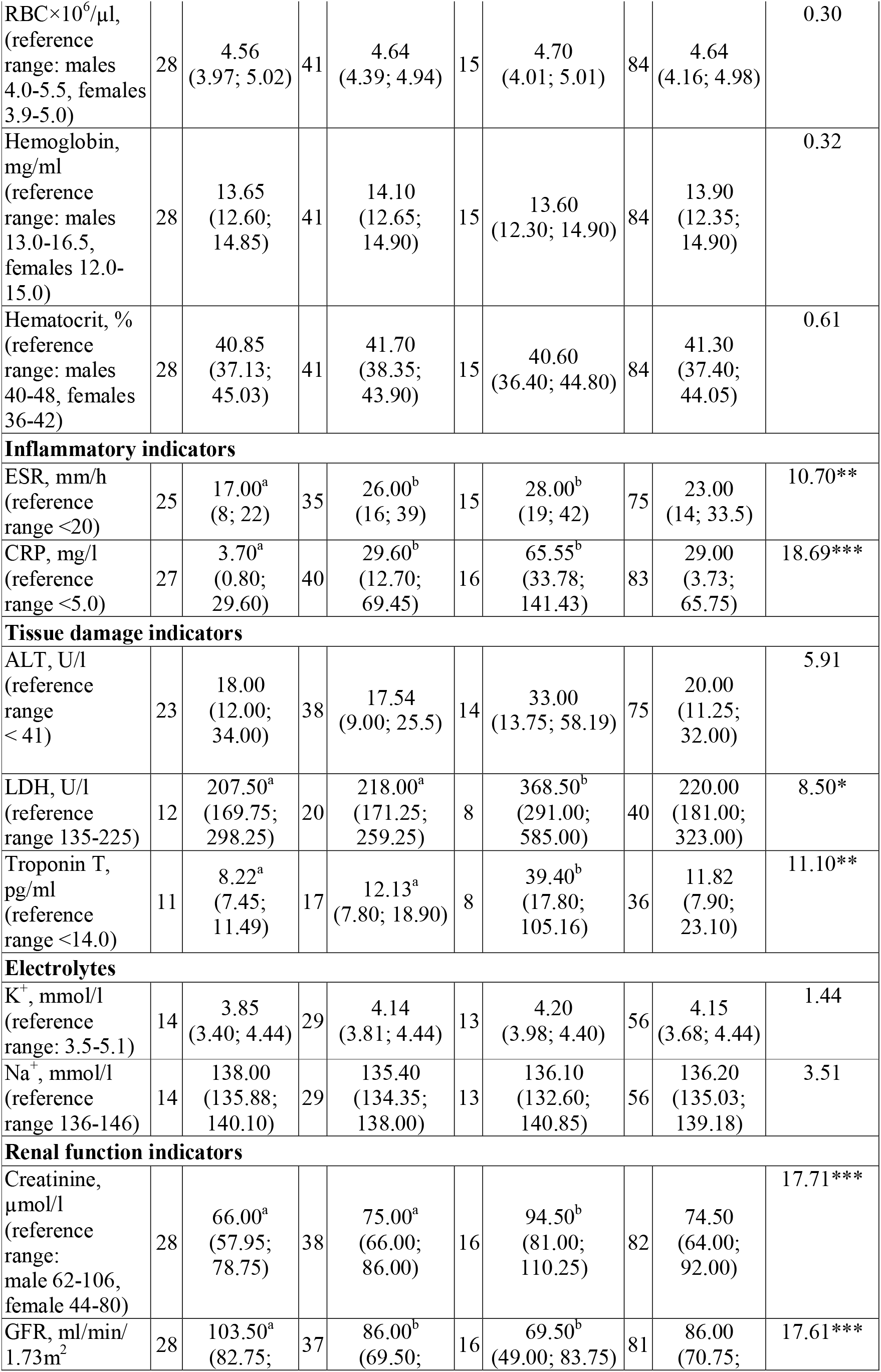
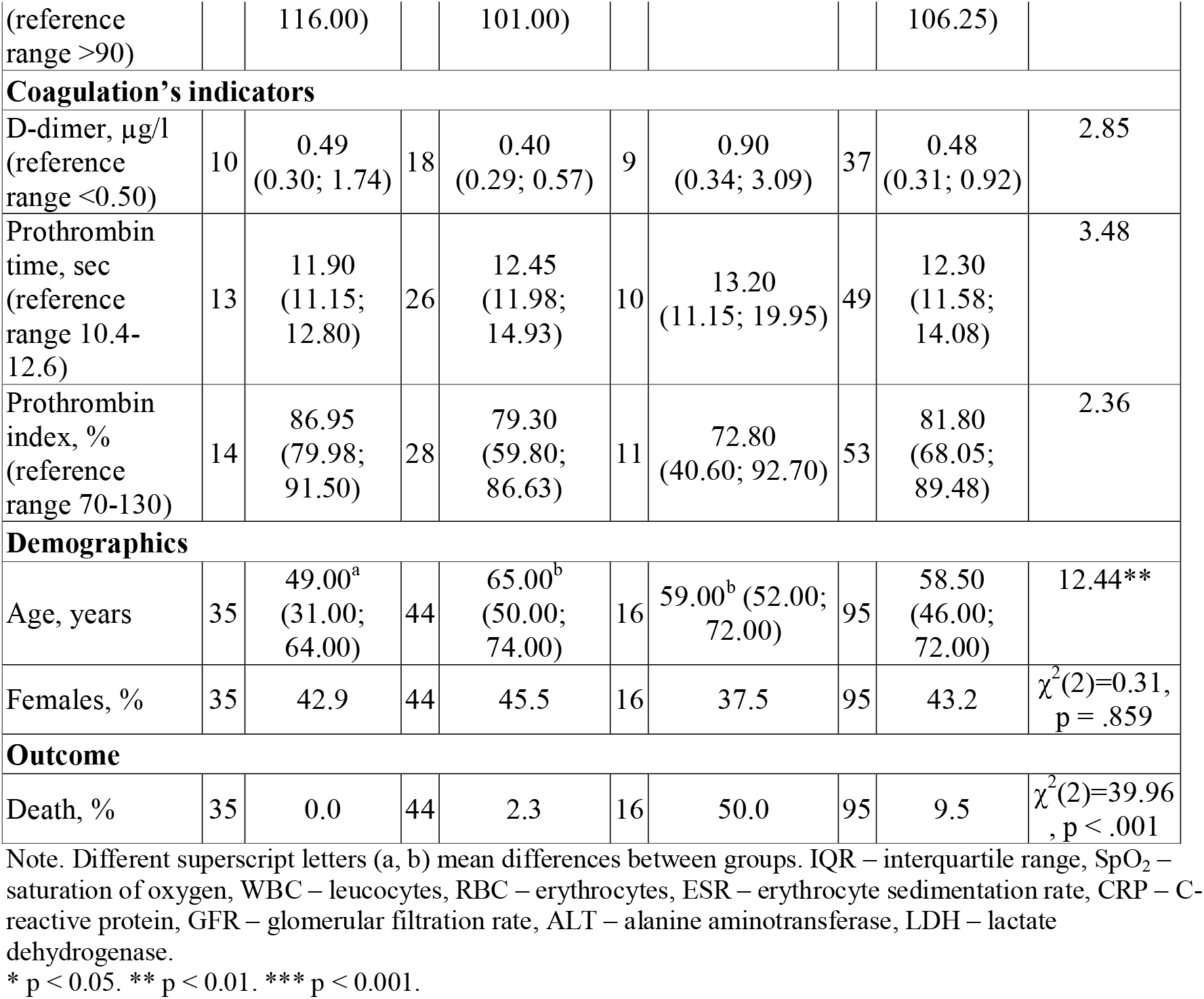
Differences in laboratory indicators, outcome, and demography in patients with mild, moderate, and severe COVID-19 disease course.

**Table 2.**
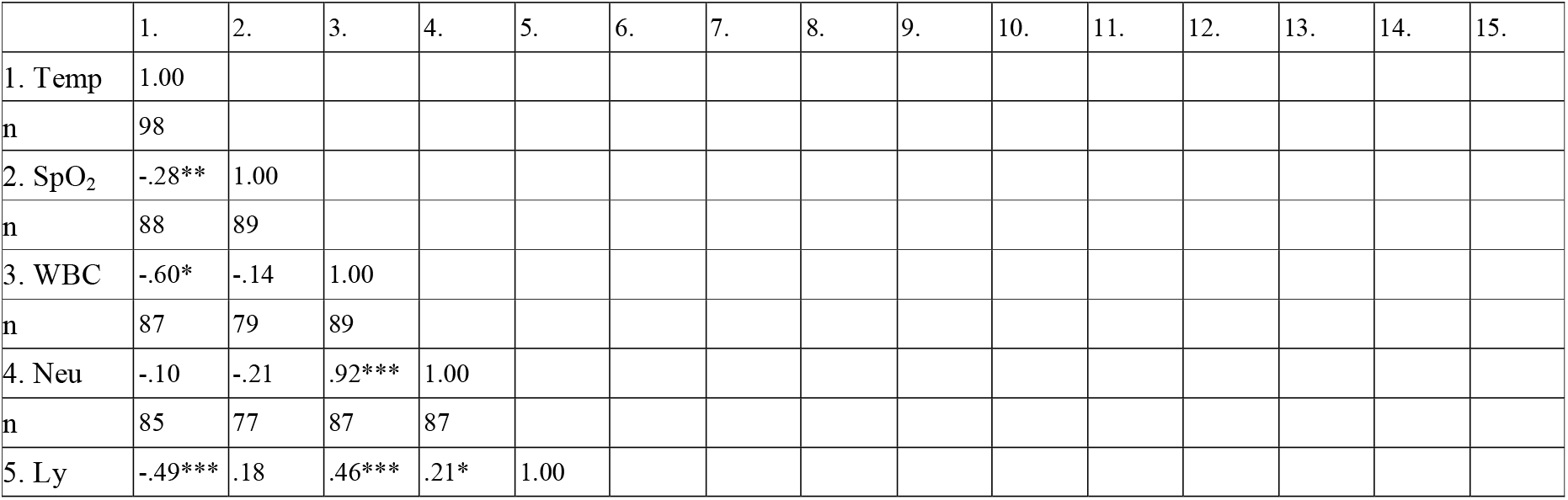
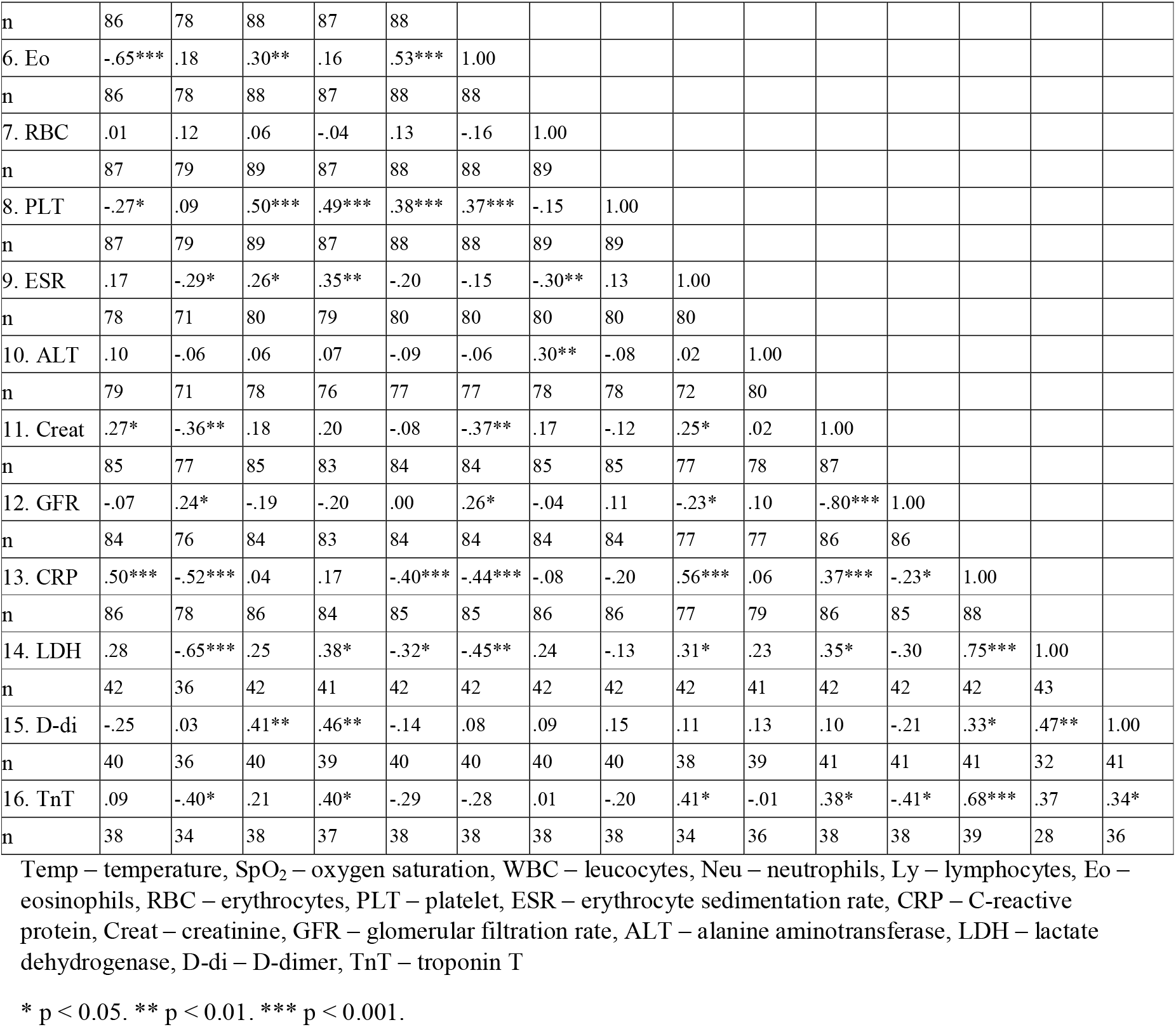
Spearman rank correlations between indicators.

### Oxygen saturation and body temperature

The level of SpO_2_ was significantly lower in the group with severe disease course than in groups with mild or moderate disease course. Patients’ temperature was significantly lower in the group with mild disease course than in the group with severe disease course.

### Blood cells

Differences in blood cells’ count were found in eosinophil count. The level of eosinophils was significantly lower in groups with moderate or severe disease course than in the group with mild disease course. There were no differences in neutrophil count, lymphocytes, monocytes, platelets, erythrocytes, hematocrit, and hemoglobin. Leukocytosis on admission was found in 11% of patients with mild COVID-19, 17% of moderate cases, and 33% of patients with severe disease course. There were no significant differences in this distribution, χ^2^(2) = 3.44, p = 0.179. Lymphocytopenia on admission was present in 54% of patients. It was observed in 39% of patients with mild disease course, in 60% of patients with moderate, and in 67% of patients with severe course. This distribution was also without significant differences, χ^2^(2) = 3.99, p = 0.136. Eosinopenia was observed in 76% of patients with COVID-19. It was distributed with significant differences among groups, χ^2^(2) = 11.94, p = 0.009. In patients with severe disease course, eosinopenia was present in 93% of cases, in patients with a moderate disease course in 85%, and in mild patients in 54%.

### Inflammatory indicators

ESR was higher in the group with moderate or severe disease course than in the group with mild disease course. The level of C-reactive protein also was higher in groups with moderate or severe disease course than in the group with mild disease course.

### Renal function indicators

The level of creatinine was significantly higher in the group with severe disease course than in groups with mild or moderate disease course. The level of GFR was significantly lower in groups with moderate or severe disease course than in the group with mild disease course.

### Tissue damage indicators

The levels of LDH and troponin T were higher in the group with severe disease course than in groups with mild or moderate disease course. The level of ALT demonstrated no differences among groups.

### Coagulation tests and electrolytes

There were no differences in D-dimer, prothrombin time and index, and potassium or sodium concentration among groups of patients on their admission to the hospital.

### Relationships among indicators

Age correlated positively with ESR, r_s_(78) = 0.29, p = 0.008, creatinine, r_s_(85) = 0.31, p = 0.003, and troponin T, r_s_(37) = 0.37, p = 0.001. Negative correlations were observed between age and temperature, r_s_(96) = -0.21, p = 0.039, RBC, r_s_(87) = -0.21, p = 0.046, and GFR, r_s_(34) = -0.55, p < 0.001.

Table 2 presents the interrelations among other indicators. Temperature correlated positively with creatinine, r_s_(85) = 0.27, p < 0.05, and CRP, r_s_(86) = 0.50, p < 0.001. It also correlated negatively with SpO_2,_ r_s_(88) = -0.28, p < 0.01, leukocyte count, r_s_(87) = -0.60, p < 0.001, lymphocytes, r_s_(86) = -0.49, p < 0.001, and eosinophils, r_s_(86) = -0.65, p < 0.001. SpO_2_ correlated positively with GFR, r_s_(76) = 0.24, p < 0.05, and negatively with ESR, r_s_(71) = - 0.29, p < 0.05, creatinine, r_s_(77) = -0.36, p < 0.01, CRP, r_s_(78) = -0.52, p = 0.001, LDH, r_s_(36) = -0.65, p = 0.001, and troponin T, r_s_(34) = -0.40, p < 0.05.

The correlation analysis revealed some associations between parameters associated with the disease course. Lymphocytes correlated positively with eosinophils, r_s_(88) = 0.53, p < 0.001, leukocytes, r_s_(88) = 0.46, p < 0.001, and platelets, r_s_(89) = 0.50, p < 0.001. CRP correlated positively with ESR, r_s_(77) = 0.56, p < 0.001, creatinine, r_s_(86) = 0.37, p < 0.001, LDH, r_s_(42) = 0.75, p < 0.001, troponin T, r_s_(39) = 0.68, p < 0.001, and negatively with eosinophils, r_s_(85) = -0.44, p = 0.001, lymphocytes, r_s_(85) = -0.40, p < 0.001, and GFR, r_s_(85) = -0.23, p < 0.05. LDH correlated positively with ESR, r_s_(42) = 0.31, p < 0.05, and creatinine, r_s_(88) = 0.46, p < 0.001, and negatively with lymphocytes, r_s_(42) = -0.32, p < 0.05, and eosinophils, r_s_(42) = - 0.45, p < 0.01. Troponin T correlated positively with ESR, r_s_(34) = 0.41, p < 0.05, and creatinine, r_s_(38) = 0.38, p < 0.05, and negatively with GFR, r_s_(38) = -0.41, p < 0.05.

## Discussion

Routine blood tests can help clinicians to assess the severity of patients with COVID-19. Our results showed that some blood parameters on admission to the hospital associate with the progression of disease severity. These parameters included eosinophils, inflammatory indicators (ESR and CRP), renal function indicators (creatinine and GFR), and tissue damage markers (LDH and troponin T). The patients with more severe disease course had higher temperature, lower blood oxygen saturation, lower eosinophils count, higher ESR, higher CRP and creatinine levels, lower GFR, and higher levels of LDH and troponin T on admission to the hospital than patients with milder disease course. In general, these results concur with previous studies [1, 5, 6, 7, 8, 11]. However, some findings should be discussed in greater detail.

For the analysis of clinical symptoms on admission, we have selected a study with a similar median interval from the onset of symptoms to admission to the hospital [8]. In our study, the mean interval from the onset of symptoms to admission to the hospital was 7±5 days, which is close to 8 days (with IQR from 6 to 11), reported by Zhang et al. [8]. In our patients, the most common symptoms were fever, cough, fatigue, and dyspnea. Their ranking was close to symptoms reported by Zhang et al., but they occurred more rarely. In our sample, gastrointestinal symptoms were reported in 6% of patients vs. 40% in Zhang et al. Probably, a relatively favorable epidemiological situation in Latvia in Spring 2020 resulted in the hospitalization of patients in a better condition compared to Zhang et al. It is also confirmed by data on abnormal chest images, reported in 59% and 99% of patients in our study and study of Zhang et al., respectively. Surprisingly, loss of smell or taste was reported only in 1% percent of patients in our study. It contrasts with an overview reporting these symptoms in 27% to 98 % of patients [12].

One of the important indicators of viral infections in general and COVID-19, in particular, is a change in blood cells count. More often, this is the lymphocytopenia [1, 6, 7, 8, 13, 14]. We have found lymphocytopenia in 54% of patients on admission. It is a relatively low level, comparing with Zhang et al. (75%) [8], Wang et al. (72%) [14], and Guang et al. (83%) [7]. Moreover, in our study, lymphocyte count on admission to the hospital demonstrated no statistical differences between patients with different disease severity.

We have revealed eosinopenia as a more visible indicator of the severity than the count of lymphocytes. Our findings suggest that patients with severe disease course had a lower count of eosinophils than patients with mild disease course, as also observed in previous studies [1, 4, 7]. Eosinophils are granulocytes, which can destruct viruses by phagocytosis during the non-specific immune response phase, similar to neutrophils. Therefore, the decreasing of eosinophils on admission to the hospital can be explained by more intensive eosinophils destruction than other groups of blood cells or by impaired hemopoiesis of these cells. Simultaneously, eosinophils and lymphocytes correlated positively, and both demonstrated a negative correlation with CRP. It confirmed a tendency of decrease in eosinophils and lymphocytes, associated with the increasing inflammatory process during the COVID-19 course.

Our study also demonstrated the relationship between COVID-19 severity and reduced renal function. It contrasts with Feng et al. [1] and Yan X. et al. [11], who revealed no differences in this function. In our study, the level of creatinine was significantly higher in the group with severe disease course than in groups with mild or moderate disease course. On the one hand, there is a relationship between reduced renal function and older age. On the other hand, the level of GFR (a marker corrected to age) was also significantly lower in groups with moderate or severe disease course than in the group with mild disease course. In our patients, reduced renal function was related to low eosinophil count. It is similar to Wagner et al. findings [15] showed an association between acute kidney injury and low lymphocyte count.

CRP and ESP are markers of an inflammatory process in infectious diseases. We have found significant elevation of their levels in severe patients with COVID-19. It confirms the findings of Zhang et al. [8] and Yan X. et al. [11]. Moreover, CRP was positively associated with tissue damage markers - LDH and troponin T. Following Yan L. et al. [13], the rise in LDH level indicates an increase in the activity and extent of lung injury in critically ill patients with COVID-19. As a marker of cardiac involvement, troponin T level is also elevated in patients with severe COVID-19 [16]. In our study, both tissue damage indicators were higher in patients with severe than moderate or mild disease course. These findings confirm previous results [1, 10, 16, 17].

Unexpectedly, in our study, D-dimer demonstrated no differences between groups contrasting with previous studies [1, 4, 8, 10]. A possible explanation is related to a relatively low count of D-dimer measurements in our patients at the beginning of the pandemic in Spring 2020.

The analysis of results showed the relationships between blood oxygen saturation, as a parameter of respiratory failure, and blood tests of inflammatory processes, renal function, and tissue damage in COVID-19 patients. It indicates that CRP, ESR, LDH, troponin T, creatinine, and GFR are indicators for monitoring patients’ condition.

A relatively low number of patients in Spring 2020 was a positive trend and, simultaneously, a limitation of our study. There was a limited opportunity to predict disease severity or death because some measures were performed on a limited number of patients. Taking into account these shortcomings, we have started a prospective study in September 2020.

In conclusions, the most unexpected discrepancy with other studies was observed in the rate of gastrointestinal symptoms and self-reported loss of smell or taste on admission to the hospital. A limited number of patients reported them in our sample. The low eosinophil count was also pronounced in our patients with severe disease course, while other studies revealed a low lymphocyte count under severe COVID-19.

The progression to severe COVID-19 was associated positively with ESR, CRP, creatinine level, LDH, troponin T and negatively associated with blood oxygen saturation, eosinophils, and GFR on admission to the hospital. Therefore, patients with severe COVID-19 demonstrated lower renal function and a higher level of inflammation and tissue damage than those with a milder disease course. These indicators can be applied in the clinical assessment and monitoring of patients. Further research in Latvian patients can be useful for the specification of significant trends and correction of clinical guidelines.

## Data Availability

Ludmila Viksna; Oksana Kolesova; Aleksandrs Kolesovs; Ieva Vanaga; Seda Arutjunana, 2020, "Clinical characteristics of COVID-19 patients (Latvia, Spring 2020)", https://doi.org/10.25143/FK2/HNMLHH, Dataverse, V1, UNF:6:bVcJ8f7pU73amPxnzHW+ZQ== [fileUNF]

## Acknowledgments

This research is funded by the Ministry of Education and Science, Republic of Latvia, project “Clinical, biochemical, immunogenetic paradigms of Covid-19 infection and their correlation with socio-demographic, etiological, pathogenetic, diagnostic, therapeutically and prognostically important factors to be included in guidelines”, project No. VPP-COVID-2020/1-0023.”

The authors would like to thank Riga Stradinš University and Riga East Clinical University Hospital for organizational support.

## Disclosure Statement

The authors report no conflict of interest.

## Notes

### Author Declarations

The Permission of the Central Medical Ethics Committee (protocol No. 01-29.1/2429) and Permission of the Ethics Committee of Riga Stradins University (protocol No. 6-1/07/14) were obtained for this study.

